# The statistical analysis of daily data associated with different parameters of the New Coronavirus COVID-19 pandemic in Georgia and their short-term interval prediction from September 2020 to February 2021

**DOI:** 10.1101/2021.04.01.21254448

**Authors:** Avtandil G. Amiranashvili, Ketevan R. Khazaradze, Nino D. Japaridze

## Abstract

In the autumn - winter period of 2020, very difficult situation arose in Georgia with the course of the pandemic of the New Coronavirus COVID-19. In particular, in November-December period of 2020, Georgia eight days was rank a first in the world in terms of COVID-19 infection rate per 1 million populations.

In this work results of a statistical analysis of the daily data associated with New Coronavirus COVID-19 infection of confirmed (C), recovered (R), deaths (D) and infection rate (I) cases of the population of Georgia in the period from September 01, 2020 to February 28, 2021 (for I - from December 05, 2020 to February 28, 2021) are presented. It also presents the results of the analysis of ten-day (decade) and two-week forecasting of the values of C, D and I, the information was regularly sent to the National Center for Disease Control & Public Health of Georgia and posted on the Facebook page https://www.facebook.com/Avtandil1948/.

The analysis of data is carried out with the use of the standard statistical analysis methods of random events and methods of mathematical statistics for the non-accidental time-series of observations. In particular, the following results were obtained.

Georgia’s ranking in the world for Covid-19 infection and deaths from September 1, 2020 to February 28, 2021 (per 1 million population) was determined. Georgia was in the first place: Infection - November 21, 22, 27, 28 and December 04, 05, 06, 09, 2020; Death - November 22, 2020.

A comparison between the daily mortality from Covid-19 in Georgia from September 1, 2020 to February 28, 2021 with the average daily mortality rate in 2015-2019 was made. The largest share value of D from mean death in 2015-2019 was 36.9% (19.12.2020), the smallest - 0.9% (21.09.2020, 24.09.2020 - 26.09.2020).

The statistical analysis of the daily and decade data associated with coronavirus COVID-19 pandemic of confirmed, recovered, deaths cases and infection rate of the population of Georgia are carried out. Maximum daily values of investigation parameters are following: C = 5450 (05.12.2020), R = 4599 (21.12.2020), D = 53 (19.12.2020), I = 30.1 % (05.12.2020). Maximum mean decade values of investigation parameters are following: C = 4337 (1 Decade of December 2020), R = 3605 (3 Decade of November 2020), D = 44 (2 Decade of December 2020), I = 26.8 % (1 Decade of December 2020).

It was found that the regression equations for the time variability of the daily values of C, R and D have the form of a tenth order polynomial.

Mean values of speed of change of confirmed -V(C), recovered - V(R) and deaths - V(D) coronavirus-related cases in different decades of months from September 2020 to February 2021 were determined. Maximum mean decade values of investigation parameters are following: V(C) = +104 cases/day (1 Decade of November 2020), V(R) = +94 cases/day (3 Decade of October and 1 Decade of November 2020), V(D) = +0.9 cases/day (1 Decade of November 2020).

Cross-correlations analysis between confirmed COVID-19 cases with recovered and deaths cases from 05.12.2020 to 28.02.2021 is carried out. So, the maximum effect of recovery is observed 13-14 days after infection, and deaths - after 13-14 and 17-18 days.

The scale of comparing real data with the predicted ones and assessing the stability of the time series of observations in the forecast period in relation to the pre-predicted one was offered.

Comparison of real and calculated predictions data of C (23.09.2020-28.02.2021), D (01.01.2021-28.02.2021) and I (01.02.2021-28.02.2021) in Georgia are carried out. It was found that daily, mean decade and two-week real values of C, D and I practically falls into the 67% - 99.99% confidence interval of these predicted values for the specified time periods (except the forecast of C for 13.10.2020-22.10.2020, when a nonlinear process of growth of C values was observed and its real values have exceeded 99.99% of the upper level of the confidence interval of forecast).

Alarming deterioration with the spread of coronavirus parameters may arise when their daily values are higher 99.99% of upper level of the forecast confidence interval. Excellent improvement - when these daily values are below 99.99% of the lower level of the forecast confidence interval.

The lockdown introduced in Georgia on November 28, 2020 brought positive results. There are clearly positive tendencies in the spread of COVID-19 to February 2021.

## 1. Introduction

More than a year has passed since the outbreak of the novel coronavirus (COVID-19) epidemic in China, recognized on March 11, 2020, as a pandemic due to its rapid spread in the world [1]. Despite the measures taken, the overall level of morbidity and mortality remains quite high. Although the main burden in the fight against the pandemic falls on epidemiologists, together with them scientists and specialists from various disciplines from around the world have joined in intensive research into this unprecedented phenomenon (including Georgia [2-6]).

Experts in the field of physical and mathematical sciences, together with doctors, make an important contribution to research on the spread of the new coronavirus COVID-19. Although scientists deal with extremely unstable time series, which greatly complicates their systematization and forecasting, various mathematical and statistical space-time models of the spread of the new coronavirus have been proposed [7-28].

Although Georgia is serious about the fight against coronavirus and it is closely monitoring the trends of the pandemic’s variability in the world and especially in neighboring countries (Armenia, Azerbaijan, Turkey, Russia), in the autumn - winter period of 2020, in this country arose very difficult situation with the course of the pandemic of COVID-19. In particular, in November-December period of 2020, Georgia eight days was rank a first in the world in terms of COVID-19 infection rate per 1 million populations. The Georgian government was forced to introduce a lockdown on November 28, 2020.

This work is a continuation of the researches [6,7].

In this work results of a statistical analysis of the daily data associated with New Coronavirus COVID-19 infection of confirmed (C), recovered (R), deaths (D) and infection rate (I) cases of the population of Georgia in the period from September 01, 2020 to February 28, 2021 (for I - from December 05, 2020 to February 28, 2021) are presented. It also presents the results of the analysis of ten-day (decade) and two-week forecasting of the values of C, D and I. The information was regularly sent to the National Center for Disease Control & Public Health of Georgia and posted on the Facebook page https://www.facebook.com/Avtandil1948/.

We used standard methods of statistical analysis of random events and methods of mathematical statistics for non-random time series of observations [6, 7, 29, 30].

## 2. Study areas, material and methods

The study area: Georgia. Data of John Hopkins COVID-19 Time Series Historical Data (with US State and County data) [https://www.soothsawyer.com/john-hopkins-time-series-data-with-us-state-and-county-city-detail-historical/; https://data.humdata.org/dataset/total-covid-19-tests-performed-by-country] and https://stopcov.ge about daily values of confirmed, recovered, deaths and infection rate coronavirus-related cases, from March 15, 2020 to February 28, 2021 are used. Detailed statistical analysis of the data was carried out for the period from September 01, 2020 to February 28, 2021. The work also used data of National Statistics Office of Georgia (Geostat) on the average monthly total mortality in Georgia in September-February 2015-2019 [https://www.geostat.ge/en/]. In the proposed work the analysis of data is carried out with the use of the standard statistical analysis methods of random events and methods of mathematical statistics for the non-accidental time-series of observations [7, 29, 30].

The following designations will be used below: Mean – average values; Min – minimal values; Max - maximal values; Range – Max-Min; St Dev - standard deviation; σ_m_ - standard error; C_V_ = 100·St Dev/Mean – coefficient of variation, %; R^2^ – coefficient of determination; r – coefficient of linear correlation; CR – coefficient of cross correlation; Lag = 1, 2…30 Day; K_DW_ – Durbin-Watson statistic; Calc – calculated data; Real - measured data; D1, D2, D3 – numbers of the month decades; α - the level of significance; C, R, D - daily values of confirmed, recovered and deaths coronavirus-related cases; V(C), V(R) and V(D) - daily values of speed of change of confirmed, recovered and deaths coronavirus-related cases (cases/day); I - daily values of infection rate coronavirus-related cases (100· C/number of coronavirus tests performed); DC – deaths coefficient, % = (100· D/C). Official data on number of coronavirus tests performed are published from December 05, 2020 [https://stopcov.ge].

The statistical programs Data Fit 7, Mesosaur and Excel 16 were used for calculations.

The curve of trend is equation of the regression of the connection of the investigated parameter with the time at the significant value of the determination coefficient and such values of K_DW_, where the residual values are accidental. If the residual values are not accidental the connection of the investigated parameter with the time we will consider as simply regression.

The calculation of the interval prognostic values of C, D and I taking into account the periodicity in the time-series of observations was carried out using Excel 16 (the calculate methodology was description in [7]). The duration of time series of observations for calculating ten-day and two-week forecasts of the values of C, D and I was 35 - 92 days.

67%…99.99%_Low - 67% 99.99% lower level of confidence interval of prediction values of C, D and I; 67%…99.99%_Upp - 67% 99.99% upper level of confidence interval of prediction values of C, D and I. In the Table 1 the scale of comparing real data with the predicted ones and assessing the stability of the time series of observations in the forecast period in relation to the pre-predicted one (period for prediction calculating) is presented.

**Table 1.** **The scale of comparing real data with the predicted ones and assessing the stability of the time series of observations in the forecast period in relation to the pre-predicted one.**

| Range of Forecast Level | Change in the forecast state in relation to the pre-predicted one |
| --- | --- |
| 67% Low ≤ Real Data ≤ 67% Upp | Preservation, stability of a time-series of observations |
| 67% Upp $<$ Real Data ≤ 95% Upp | Noticeable deterioration, stability of a time-series of observations |
| 95% Upp $<$ Real Data ≤ 99% Upp | Significant deterioration, stability of a time-series of observations |
| 99% Upp $<$ Real Data ≤ 99.99% Upp | Sharp deterioration, stability of a time-series of observations |
| Real Data $>$ 99.99% Upp | Alarming deterioration, violation of the stability of a time-series of observations |
| 95% Low ≤ Real Data $<$ 67% Low | Noticeable improvement, stability of a time-series of observations |
| 99% Low ≤ Real Data $<$ 95% Low | Significant improvement, stability of a time-series of observations |
| 99.99% Low ≤ Real Data $<$ 99% Low | Sharp improvement, stability of a time-series of observations |
| Real Data $<$ 99.99% Low | Excellent improvement, violation of the stability of a time-series of observations |

## 3. Results and Discussion

The results in the Fig. 1-20 and Table 2-4 are presented.

**Table 2.** Statistical characteristics of the daily data associated with New Coronavirus COVID-19 pandemic of confirmed, recovered, deaths cases and infection rate of the population of Georgia.

| Parameter | Confirmed | Recovered | Deaths | Infection Rate, % |
| --- | --- | --- | --- | --- |
| Period | 01.09.2020-28.02.2021 |  |  | 05.12.2020-28.02.2021 |
| Mean | 1488 | 1458 | 19 | 9.7 |
| Min | 20 | 0 | 0 | 0.9 |
| Max | 5450 | 4599 | 53 | 30.1 |
| Range | 5430 | 4599 | 53 | 29.2 |
| St Dev | 1385 | 1383 | 15 | 7.6 |
| $\sigma_m$ | 103 | 103 | 1.1 | 0.8 |
| $C_V$ (%) | 93.1 | 94.8 | 77.9 | 78.3 |

**Fig.1.**
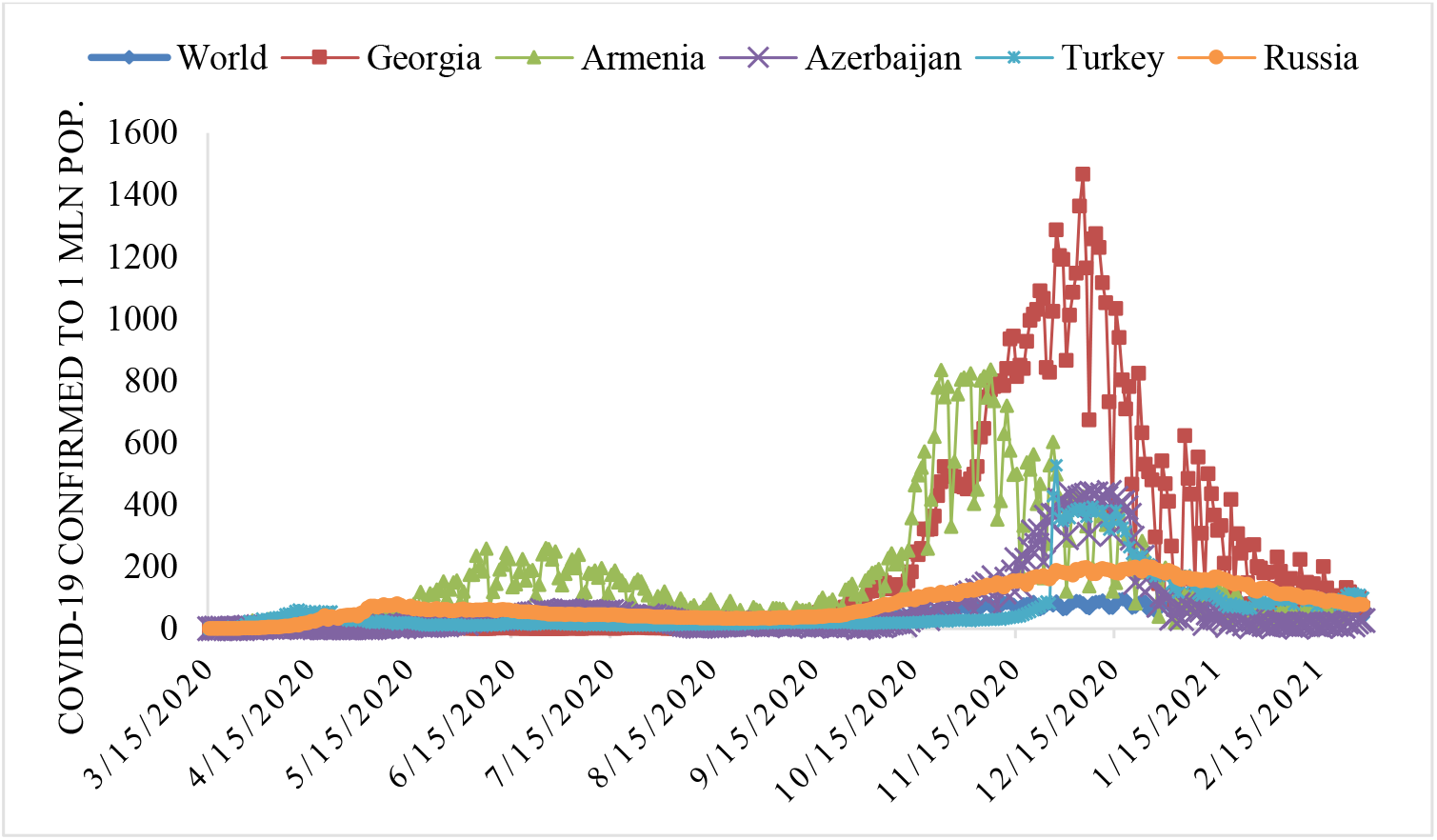
Time-series of Covid-19 confirmed cases to 1 million populations in World, Georgia and its neighboring countries from March 15, 2020 to February 28, 2021 (excluding data for Turkey on 10.12.2020 as incorrect).

### 3.1 Comparison of time-series of Covid-19 confirmed and deaths cases in World, Georgia and its neighboring countries from March 15, 2020 to February 28, 2021

Time-series of Covid-19 confirmed and deaths cases (to 1 million populations) in World, Georgia and its neighboring countries from March 15, 2020 to February 28, 2021 in Fig. 1 and 2 are presented.

**Fig.2.**
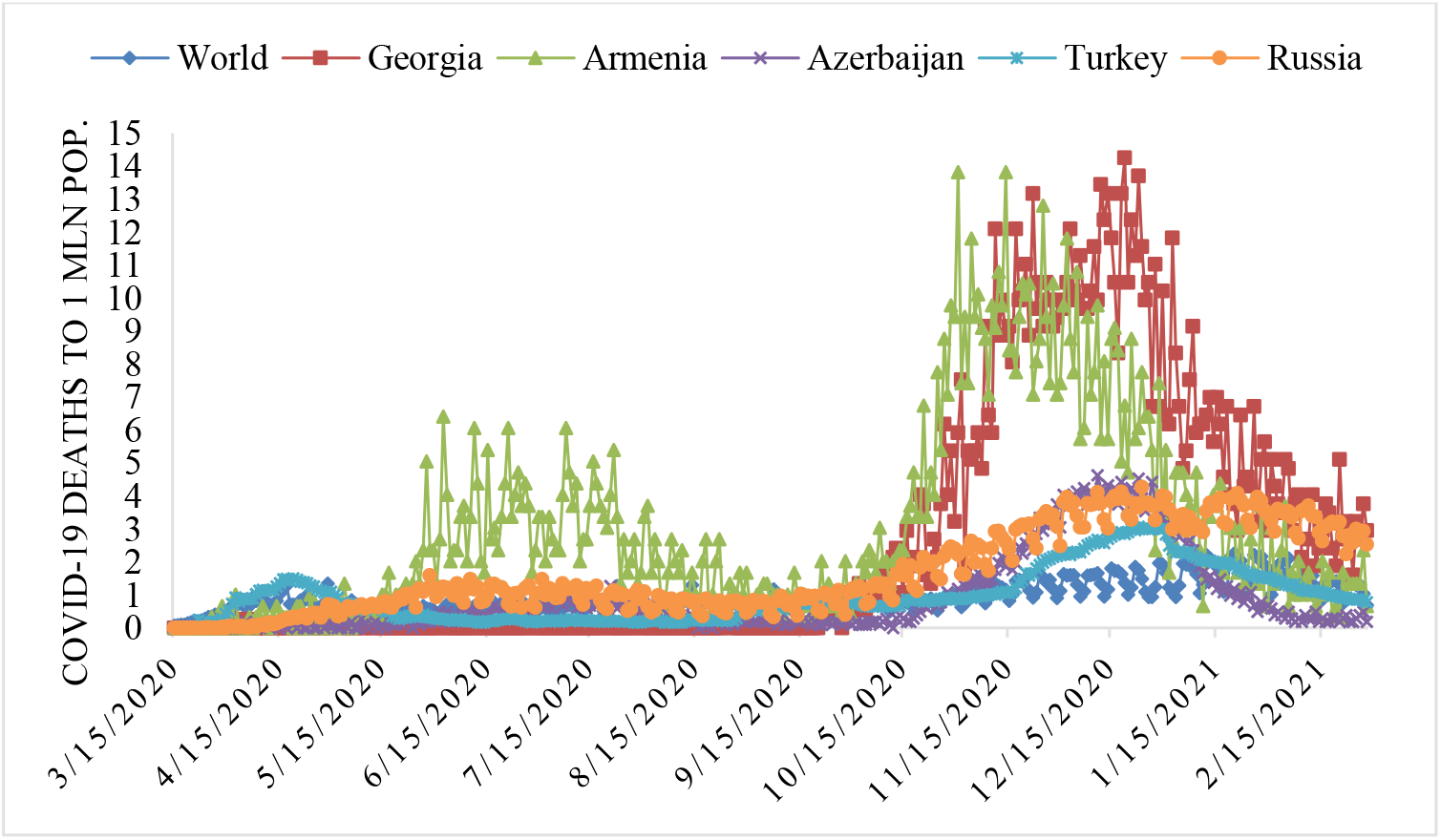
Time-series of deaths cases from Covid-19 to 1 million populations in the world, Georgia and its neighboring countries from March 15, 2020 to February 28, 2021.

Dynamics of time variability of the values of C per 1 million populations is as follows (Fig. 1):

- World. Almost bimodal distribution with a max on January 7, 2021 (C = 113). Mean value of C = 41.
- Georgia. Bimodal distribution with a max on December 5, 2021 (C = 1466). Mean value of C = 208.
- Armenia. Two modal distribution. The first extremum was on June 25, 2020 (C = 260), the second - on November 7, 2020 (C =836). Mean value of C = 165.
- Azerbaijan. Two modal distribution. The first extremums were on June 24, 2020 and July 1, 2020 (C = 58), the second - on December 13, 2020 (C = 439). Mean value of C = 66.
- Turkey. Two modal distribution. The first extremum was on April 11, 2020 (C = 61), the second - on November 11, 2020 (C = 528). Mean value of C = 64.
- Russia. Two modal distribution. The first extremum was on May 11, 2020 (C = 80), the second – on December 24, 2020 (C = 202). Mean value of C = 82.

Dynamics of time variability of the values of D per 1 million populations is as follows (Fig. 2):

- World. Almost bimodal distribution with a max on January 20, 2021 (D = 2.25). Mean value of D = 0.92.
- Georgia. Bimodal distribution with a max on December 19, 2021 (D = 14.26). Mean value of D = 2.69.
- Armenia. Two modal distribution. The first extremum was on June 2, 2020 (D = 6.41), the second - on October 31, 2020 and November 14, 2020 14 (D = 13.84). Mean value of D = 3.07.
- Azerbaijan. Two modal distribution. The first extremum was on July 21, 2020 (D = 1.28), the second - on December 11, 2020 (D = 4.64). Mean value of D = 0.90.
- Turkey. Two modal distribution. The first extremum was on April 19, 2020 (D = 1.51), the second - on December 23, 2020 (D =3.07). Mean value of D = 0.97.
- Russia. Two modal distribution. The first extremum was on May 29, 2020 (D = 1.59), the second - on December 24, 2020 (D = 4.28). Mean value of D = 1.65.

The mean values of DC (%) are: World – 2.23, Georgia – 1.30, Armenia – 1.86, Azerbaijan – 1.37, Turkey – 1.52, Russia – 2.02.

Thus, in the period from March 15, 2020 to February 28, 2021, Georgia ranked first in terms of the average level of infection with COVID-19 in comparison with neighboring countries, and second in terms of mortality (after Armenia).

At the same time, Georgia ranked last on the deaths coefficient from COVID-19.

In Fig. 3-5 information of the ranking and first ten ranking Georgia’s in the world for Covid-19 infection and deaths from September 1, 2020 to February 28, 2021 (per 1 million population) are presented.

**Fig.3.**
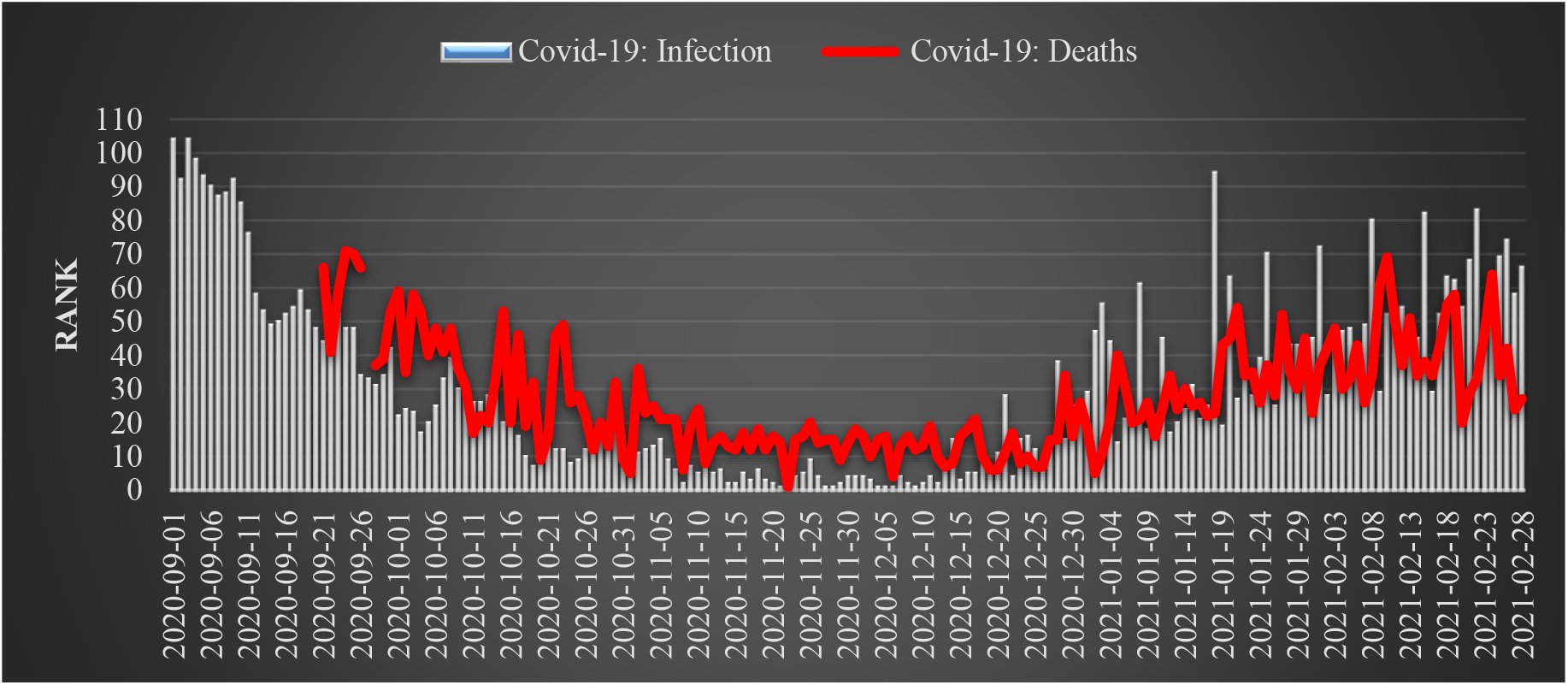
Georgia’s ranking in the world for Covid-19 infection and deaths from September 1, 2020 to February 28, 2021 (per 1 million population).

Thus, Georgia in the world in terms of infection and mortality from COVID-19 from September 2020 to February 2021 occupied the following ranks (Fig. 3).

Infection. September 2020, mean - 63, range: 104 - 31; October 2020, mean - 20, range: 39 - 7; November 2020, mean - 6, range: 15 - 1; December 2020, mean - 9, range: 38 - 1; January 2021, mean - 36, range: 94 - 14; February 2021, mean - 55, range: 83 – 28.

Deaths. September 2020, mean - 56, range: 71 - 37; October 2020, mean - 32, range: 59 - 9; November 2020, mean - 16, range: 36 - 1; December 2020, mean - 14, range: 34 - 4; January 2021, mean - 29, range: 54 - 5; February 2021, mean - 41, range: 69 – 20.

Dates of the first ten ranking Georgia’s in the world for Covid-19 infection in the studies period are following (Fig. 4):

**Fig.4.**
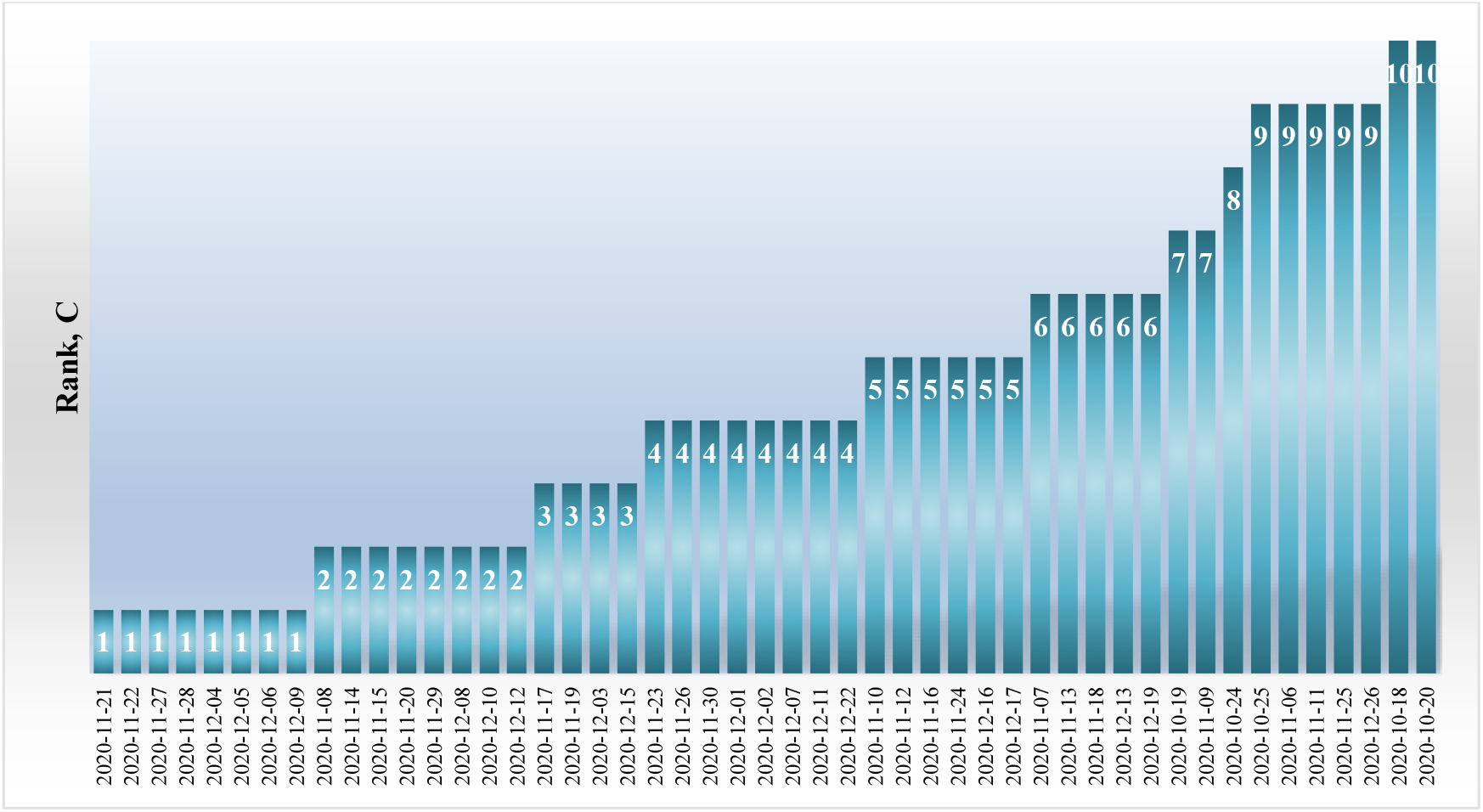
Dates of the first ten ranking Georgia’s in the world for Covid-19 infection in the studies period

Rank 1 – November 21, 22, 27, 28 and December 04, 05, 06, 09, 2020 (8 days); Rank 2 – November 8, 14, 15, 20, 29 and December 8, 10, 12, 2020 (8 days); Rank 3 - November 17, 19 and December 3, 15, 2020 (4 days); Rank 4 - November 23, 26, 30 and December 01, 02, 07, 11, 22, 2020 (8 days); Rank 5 – November 10, 12, 16, 24 and December 16, 17, 2020 (6 days); Rank 6 - November 07, 13, 18 and December 13, 19, 2020 (5 days); Rank 7 – October 19 and November 09, 2020 (2 days); Rank 8 - October 24, 2020 (1 day); Rank 9 – October 25 and November 06, 11, 25, 26, 2020 (5 days); Rank 10 - October 18, 20, 2020 (2 days).

In the first ten ranking Georgia for Covid-19 infection were in the world in October 2020 – 5 days, in November 2020 – 25 days, and in December 2020 –19 days.

Dates of the first ten ranking Georgia’s in the world for Covid-19 deaths in the studies period are following (Fig. 5): Rank 1 – November 22, 2020 (1 day); Rank 4 - December 06, 2020 (1 day); Rank 5 – November 01, 2020 and January 02, 2021 (2 day); Rank 6 - November 08 and December 19, 20, 2020 (3 days); Rank 7 - December 13, 25, 26, 2020 (3 days); Rank 8 - November 11 and December 14, 23, 2020 (3 days); Rank 9 – October 20, 31 and November 29, 2020 (3 days); Rank 10 - December 03, 12, 18, 24, 2020 (4 days).

**Fig.5.**
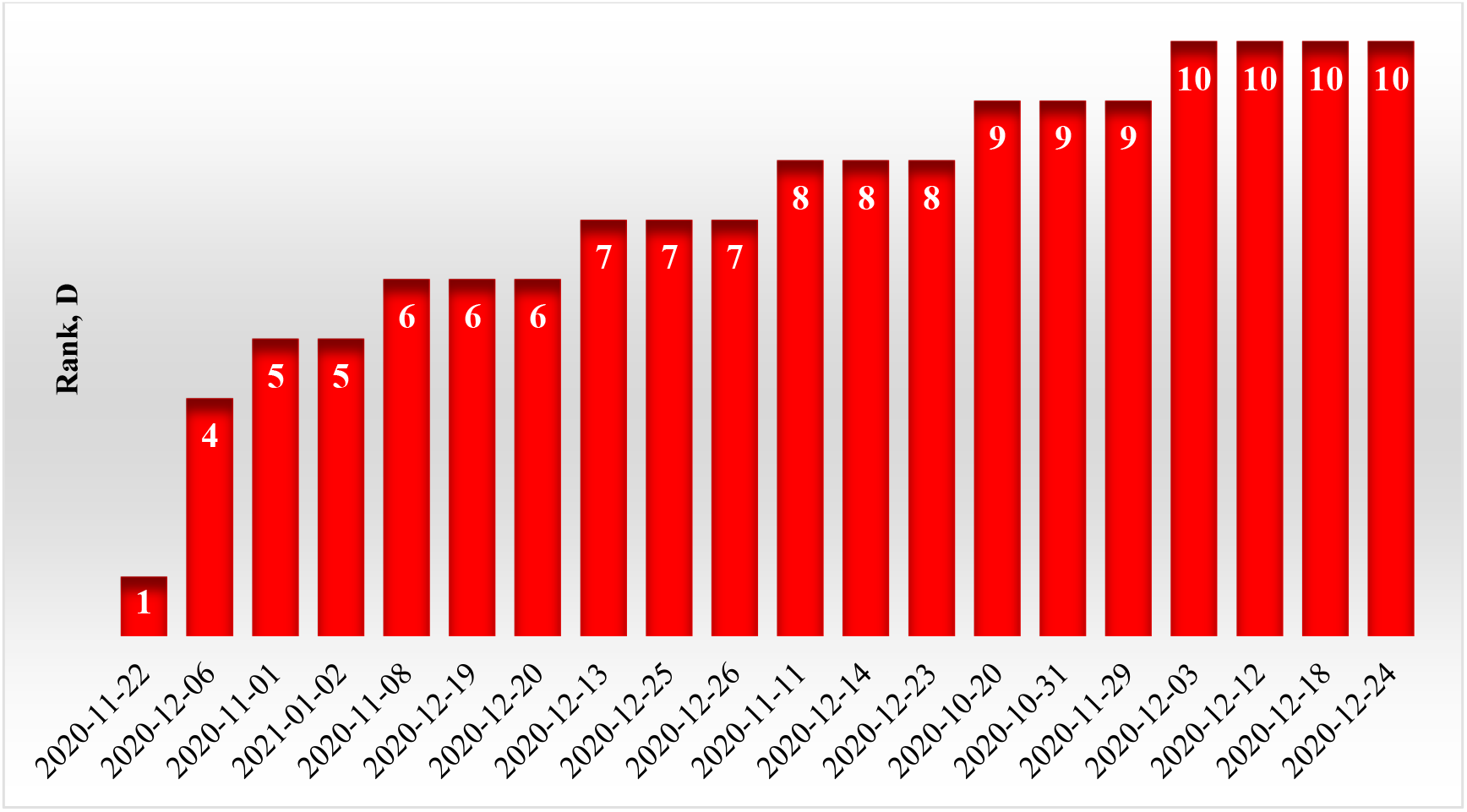
Dates of the first ten ranking Georgia’s in the world in the world for Covid-19 deaths in the studies period.

In the first ten ranking Georgia for Covid-19 deaths were in the world in October 2020 – 2 days, in November 2020 – 5 days, in December 2020 – 12 days, and in January 2021 – 1 day.

### 3.2 Comparison of daily death from Covid-19 in Georgia with daily mean death in 2015-2019 from September 1, 2020 to February 28, 2021

In Fig. 6 data of the daily death from Covid-19 in Georgia from September 1, 2020 to February 28, 2021 in comparison with daily mean sum death in 2015-2019. The daily mean sum death in 2015-2019 in different months are: September −112, October - 123, November - 135, December - 144, January - 155, February - 146.

**Fig.6.**
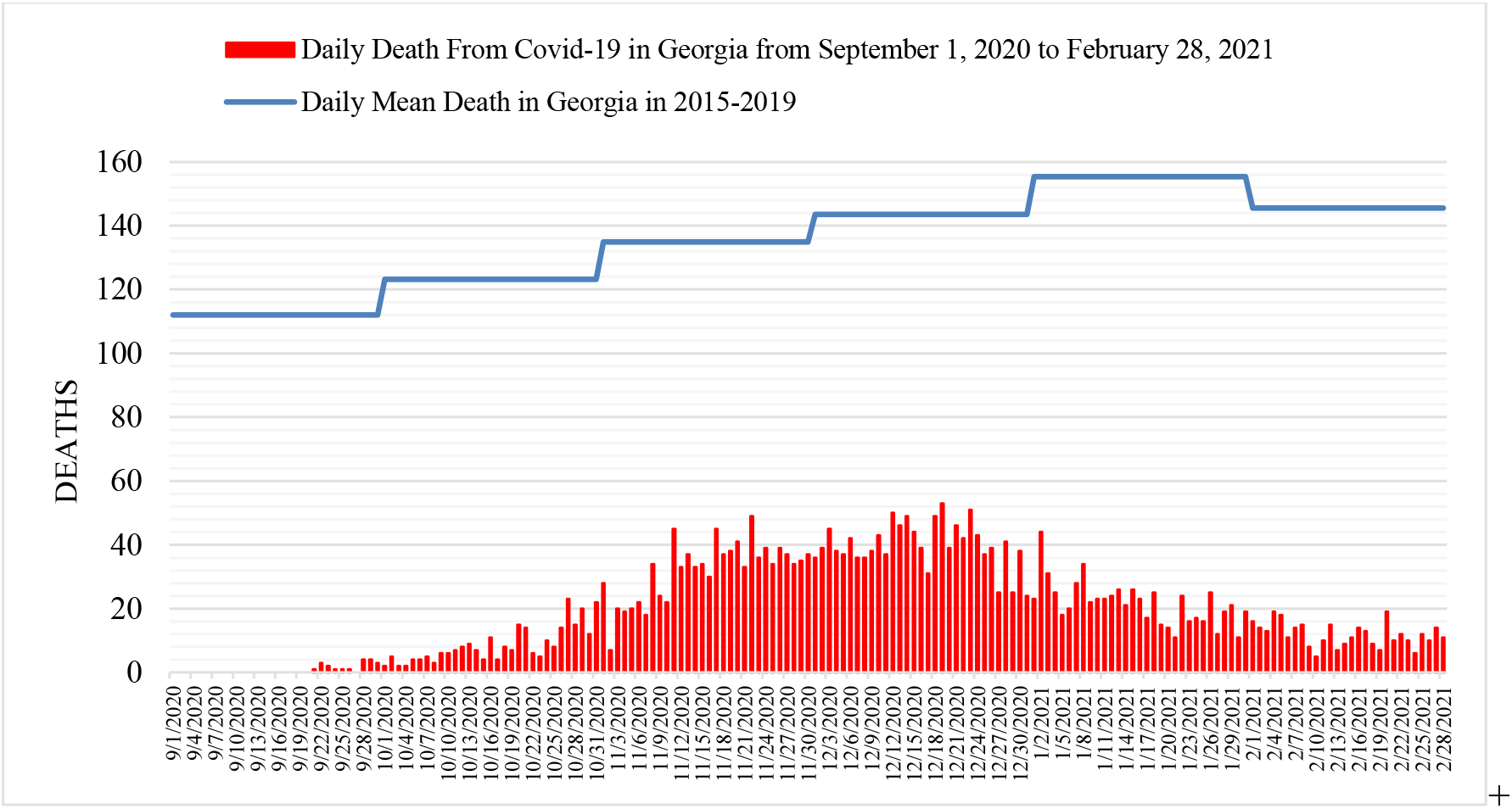
Daily death from Covid-19 in Georgia from September 1, 2020 to February 28, 2021 in comparison with daily mean death in 2015-2019.

The daily death from Covid-19 from September 1, 2020 to February 28, 2021 are: September 2020, mean - 1, range: 0 - 4; October 2020, mean - 9, range: 2 - 23; November 2020, mean - 32, range: 7 - 49; December 2020, mean - 40, range: 24 - 53; January 2021, mean - 22, range: 11 - 44; February 2021, mean - 12, range: 5 – 19.

Share value of D from mean death in 2015-2019 are (%, Fig 7.) are: September 2020, mean – 0.6, range: 0 – 3.6; October 2020, mean – 7.0, range: 1.6 – 18.7; November 2020, mean – 23.7, range: 5.2 – 36.3; December 2020, mean – 27.8, range: 16.3 – 36.9; January 2021, mean – 14.0, range: 7.1 – 28.3; February 2021, mean – 8.1, range: 3.4 – 13.1.

**Fig.7.**
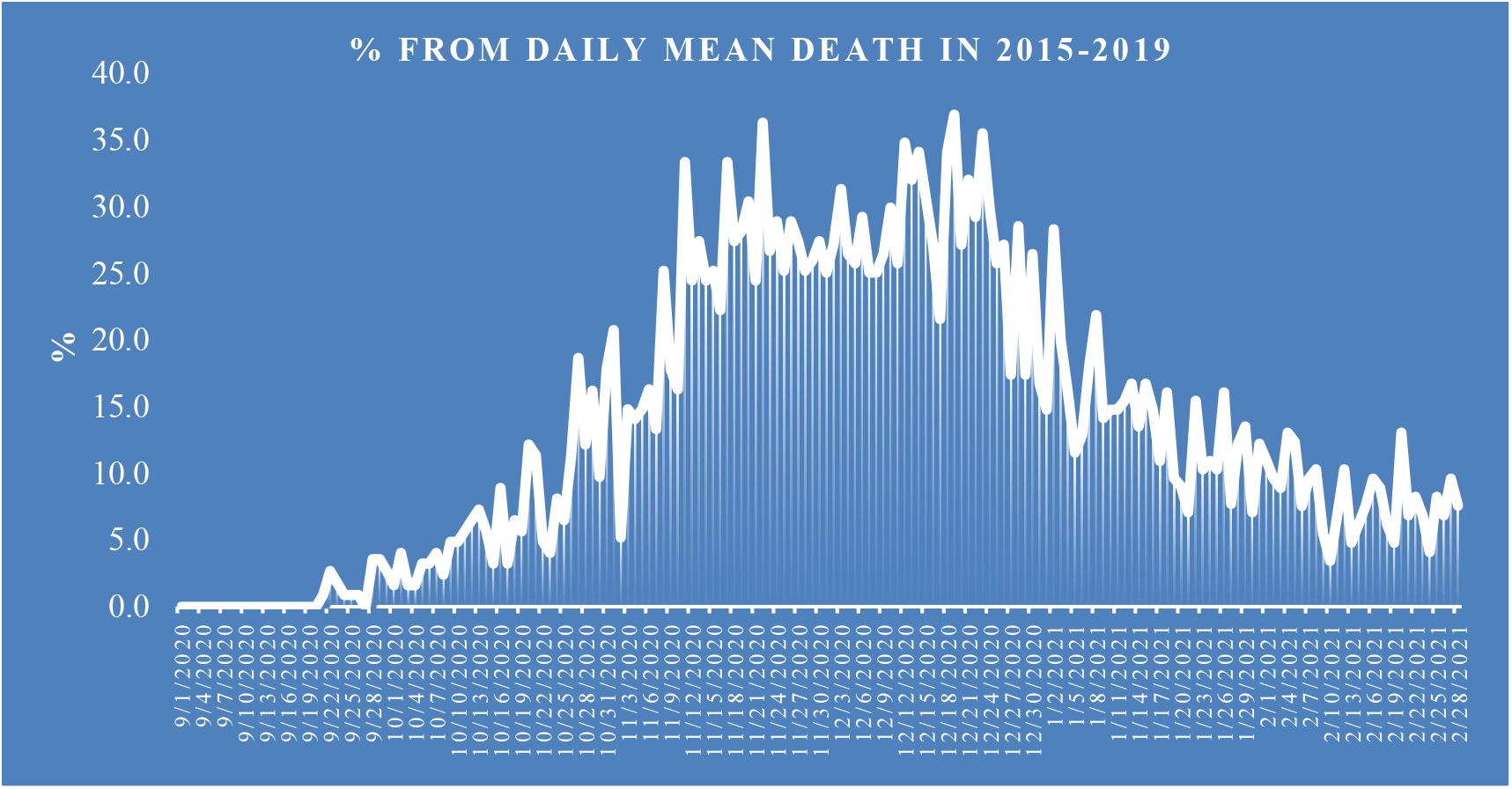
Share value of D from mean death in 2015-2019.

### 3.3 The statistical analysis of the daily and decade data associated with New Coronavirus COVID-19 pandemic

Results of the statistical analysis of the daily and decade data associated with New Coronavirus COVID-19 pandemic in Georgia from September 1, 2020 to February 28, 2021 in Tables 2,3 and Fig. 8 – 16 are presented.

**Table 3.**
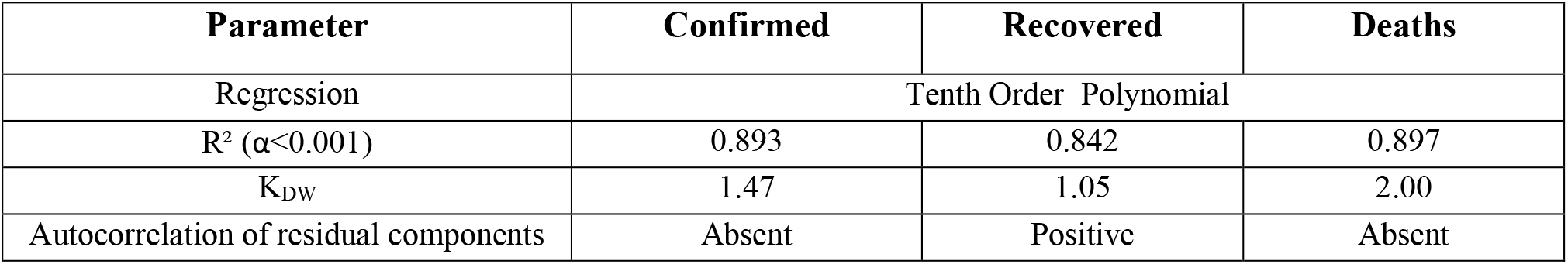
Form of the equations of the regression of the time changeability of the daily values of C, R and D from September 01, 2020 to February 28, 2021 in Georgia.

| Parameter | Confirmed | Recovered | Deaths |
| --- | --- | --- | --- |
| Regression | Tenth Order Polynomial |  |  |
| $R^2 (\alpha < 0.001)$ | 0.893 | 0.842 | 0.897 |
| $K_{DW}$ | 1.47 | 1.05 | 2.00 |
| Autocorrelation of residual components | Absent | Positive | Absent |

**Fig.8.**
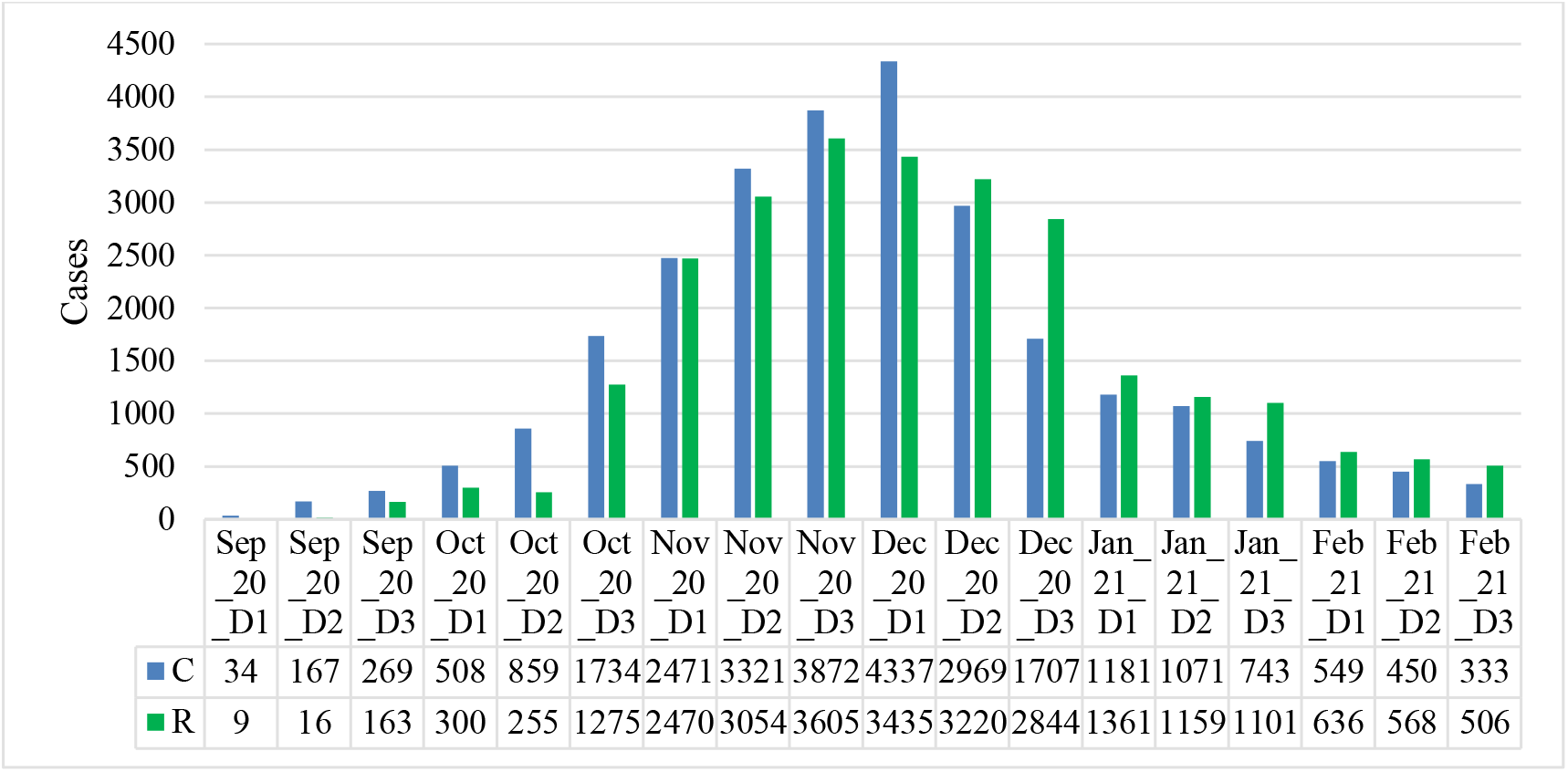
Mean values of confirmed and recovered coronavirus-related cases in different decades of months in Georgia from September 2020 to February 2021.

The mean and extreme values of the studied parameters are as follows (Table 2): C - mean - 1488, range: 20 - 5450; R - mean - 1458, range: 0 - 4599; D - mean - 19, range: 0 - 53; I (%) - mean – 9.7, range: 30.1 – 0.9. All studied parameters are subject to strong variations. 77.9 % (D) ≤C_**V**_ ≤94.8%(R).

Mean decade values of confirmed and recovered coronavirus-related cases varies within the following limits (Fig. 8): C – from 34 (1 Decade of September 2020) to 4337 (1 Decade of December 2020); R – from 9 (1 Decade of September 2020) to 3605 (3 Decade of November 2020).

In the third decade of February 2021, the values of C and R in relation to the maximum, respectively, decreased by 13 and 7 times.

Mean decade values of deaths coronavirus-related cases (Fig. 9) varies from 0 (1 and Decades of September 2020) to 44 (2 Decade of December 2020).

**Fig.9.**
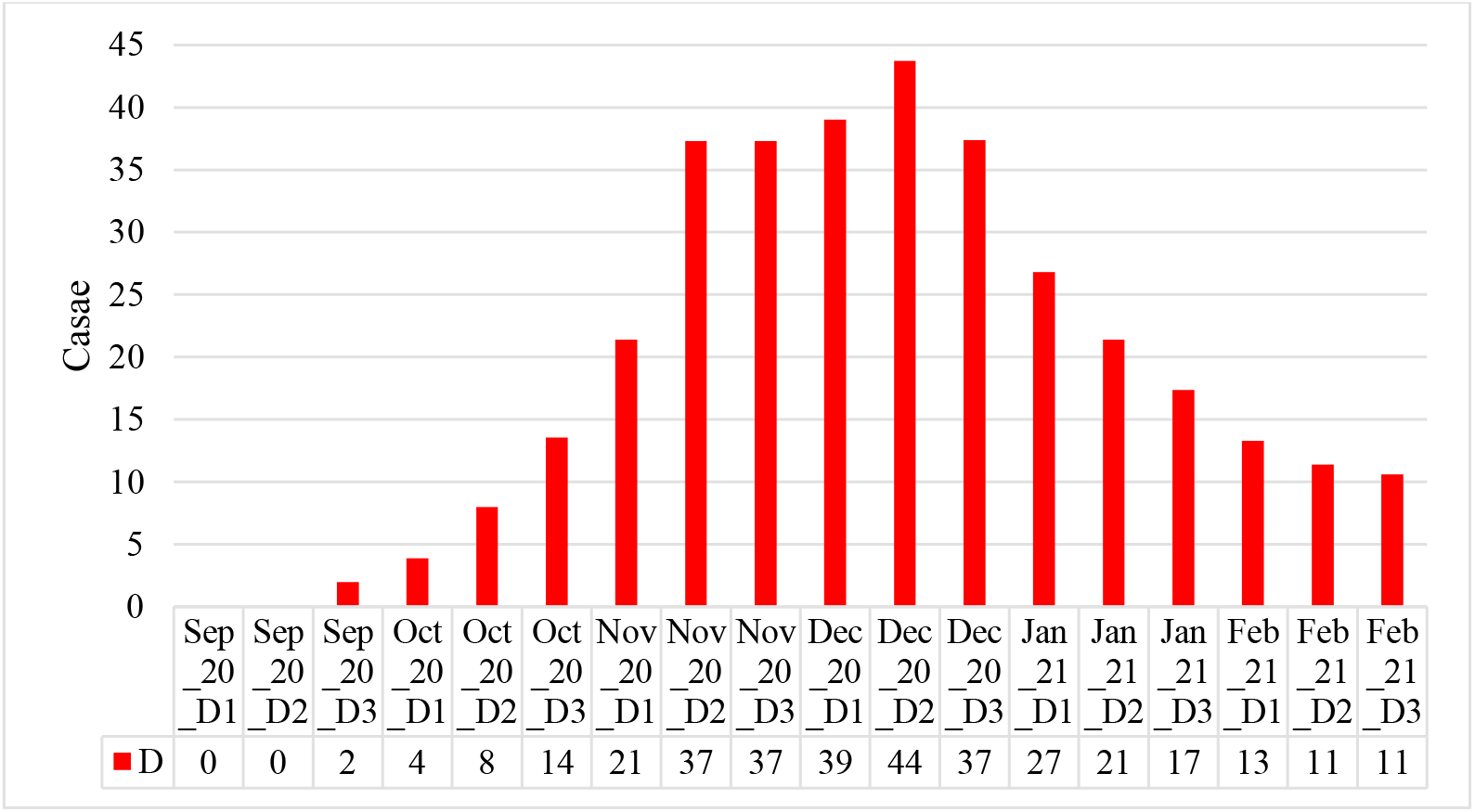
Mean values of deaths coronavirus-related cases in different decades of months in Georgia from September 2020 to February 2021.

In the second and third decades of February 2021, the values of D in relation to the maximum decreased by 4 times.

Mean decade values of infection rate coronavirus-related cases (Fig. 10) varies from 26.8% (1 and Decade of December 2020) to 1.9% (3 Decade of February 2020). In the third decades of February 2021 the values of I in relation to the maximum decreased by 14 times.

**Fig. 10.**
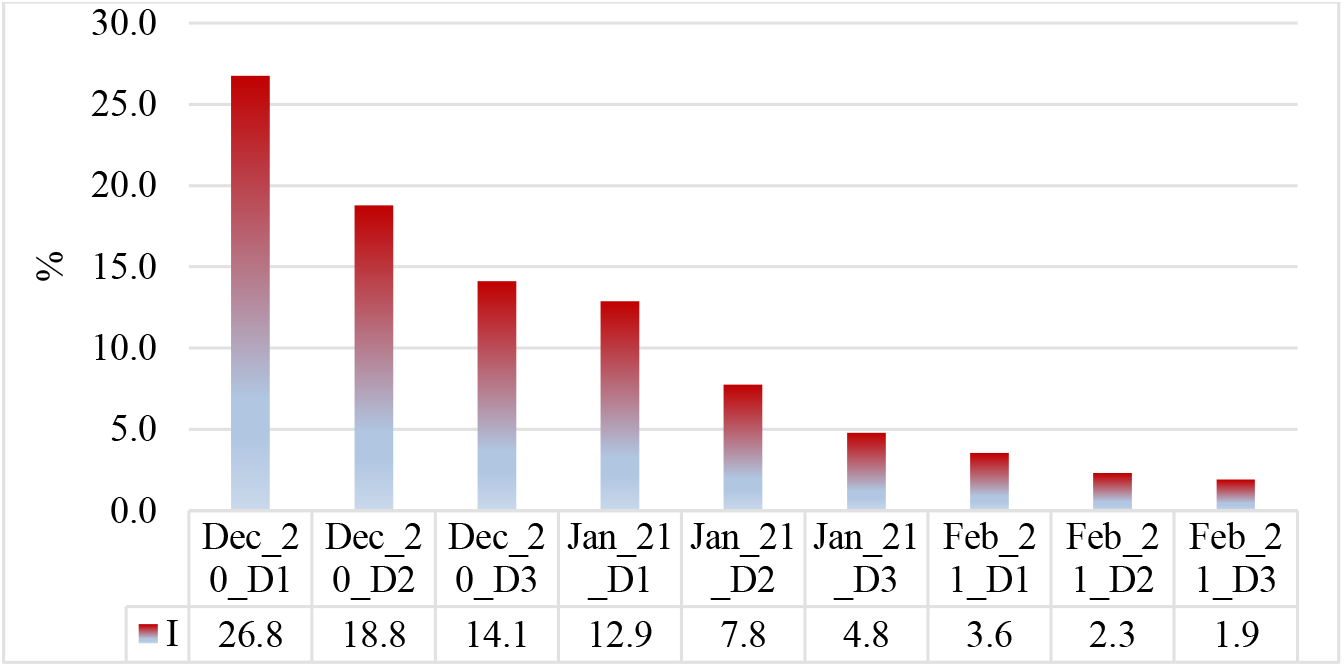
Mean values of infection rate coronavirus-related cases in different decades of months in Georgia from September 2020 to February 2021.

Time changeability of the daily values of C, R and D are satisfactorily described by the tenth order polynomial (Table 3, Fig. 11,12). For clarity, the data in Fig. 11 and 12 are presented in relative units (%) in relation to their average values.

**Fig.11.**
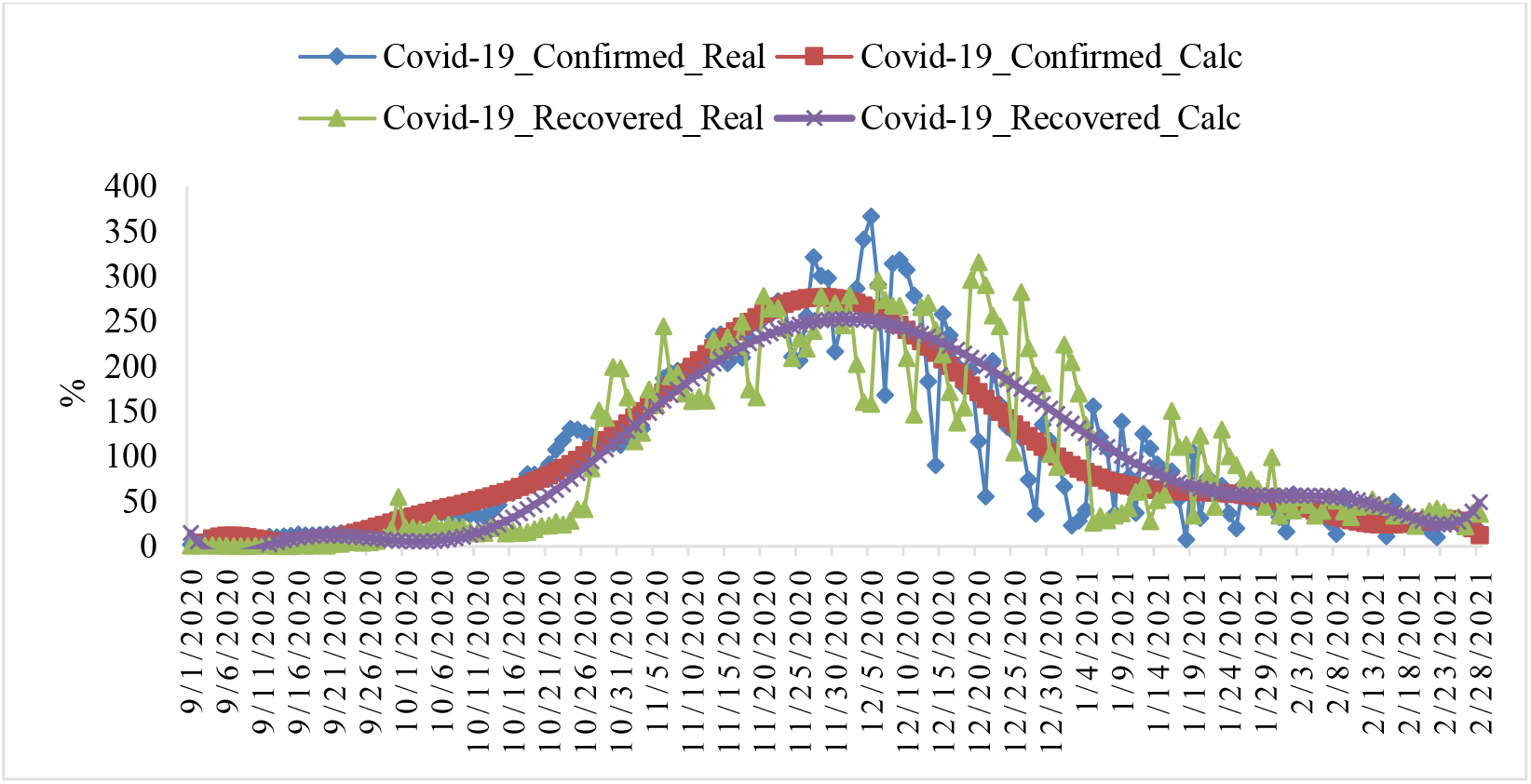
Changeability of the real and calculated daily values of coronavirus-related confirmed and recovered cases from September 01, 2020 to February 28, 2021 in Georgia (normed on mean values, %).

**Fig.12.**
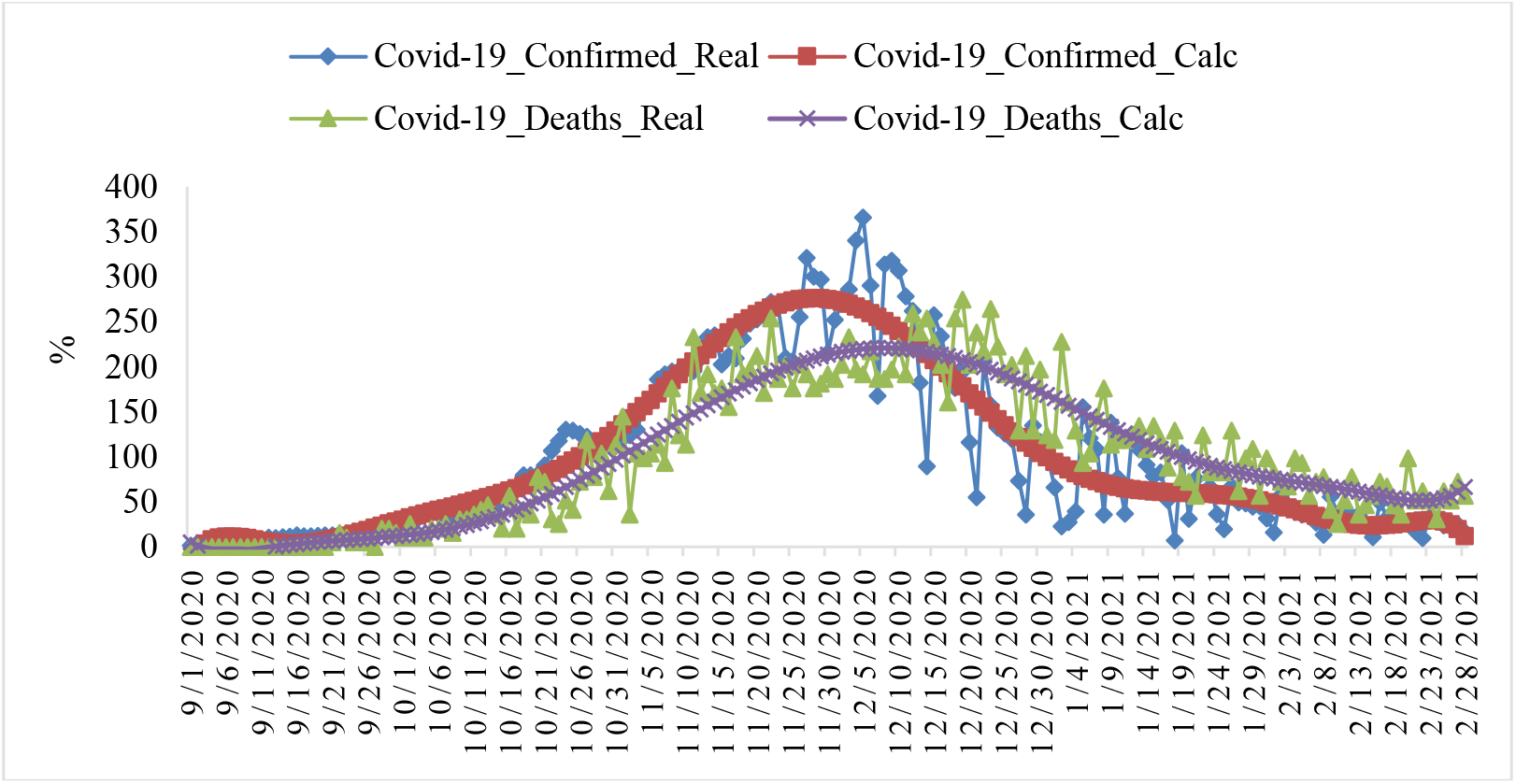
Changeability of the real and calculated daily values of coronavirus-related confirmed and deaths cases from September 01, 2020 to February 28, 2021 in Georgia (normed on mean values, %).

Note that from Fig. 11 and 12 clearly show the delay of the time series values of R and D in relation to C.

In Fig. 13,14 data about mean values of speed of change of confirmed, recovered and deaths coronavirus-related cases in different decades of months from September 2020 to February 2021 were presented. Max mean decade values of investigation parameters are following: V(C) = +104 cases/day (1 Decade of November 2020), V(R) = +94 cases/day (3 Decade of October and 1 Decade of November 2020), V(D) = +0.9 cases/day (1 Decade of November 2020). Min mean decade values of investigation parameters are following: V(C) = −104 cases/day (2 Decade of December 2020), V(R) = −77 cases/day (3 Decade of December 2020), V(D) = −0.8 cases/day (1 Decade of January 2020).

**Fig.13.**
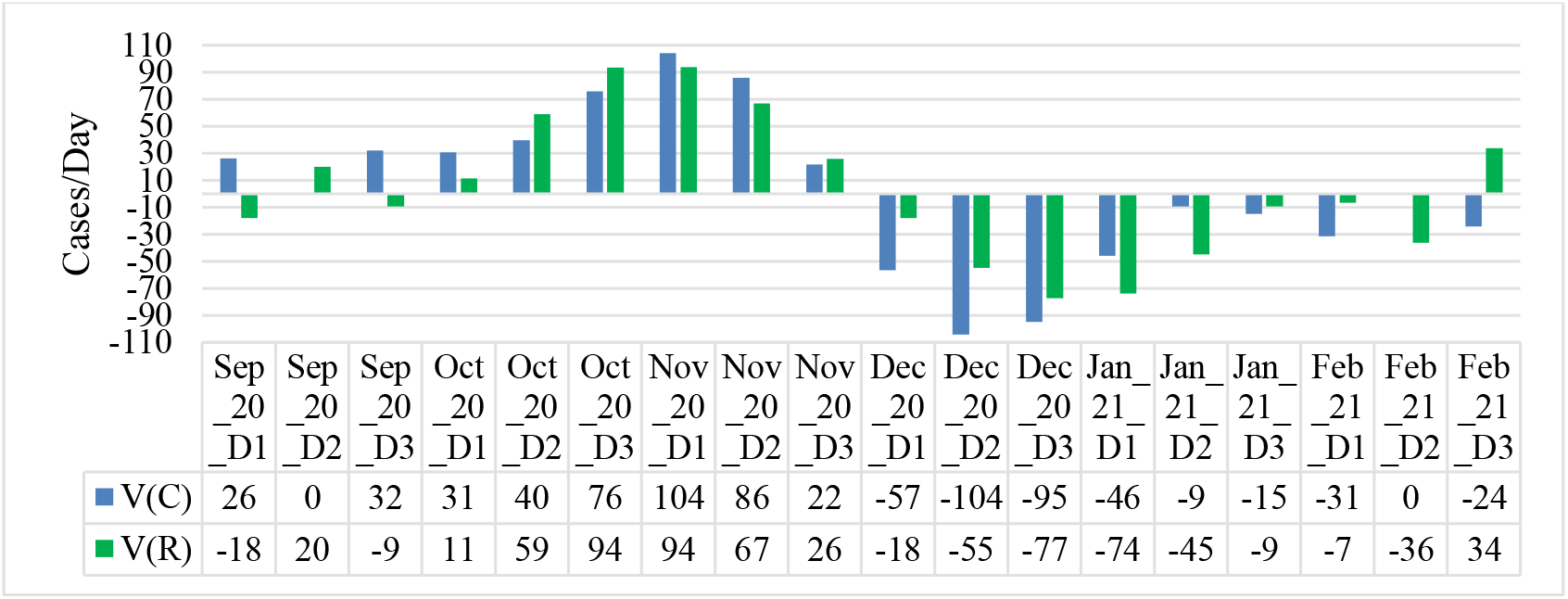
Mean values of speed of change of confirmed and recovered coronavirus-related cases in different decades of months in Georgia from September 2020 to February 2021.

**Fig.14.**
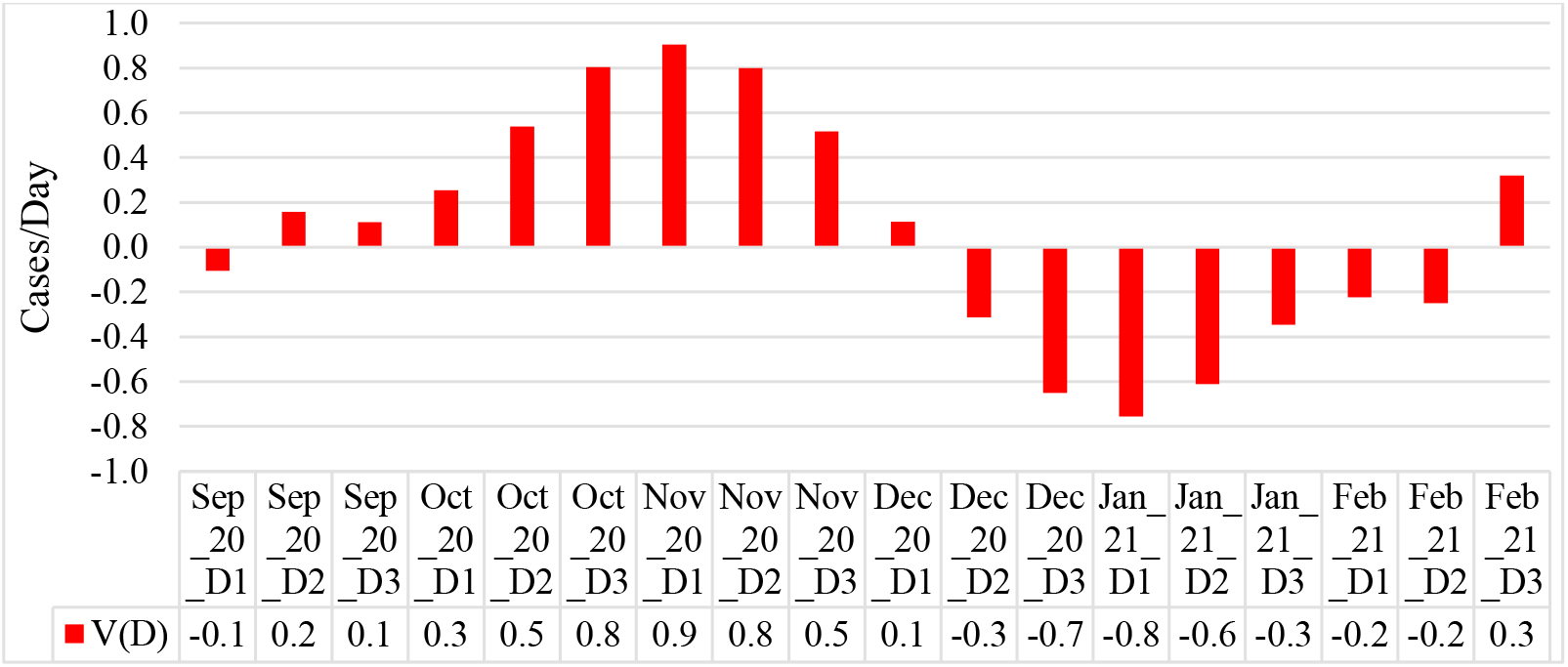
Mean values of speed of change of deaths coronavirus-related cases in different decades of months in Georgia from September 2020 to February 2021.

Thus, as follows from the above data, the lockdown introduced in Georgia on November 28, 2020 brought positive results. Already in February 2021, there was a significant decrease in the values of the studied parameters associated with New Coronavirus COVID-19.

This fact is also confirmed by the data shown in Fig. 15, in which linear correlation and regression between the confirmed cases of COVID-19 and number of tests performed in Georgia in different months are presented.

**Fig.15.**
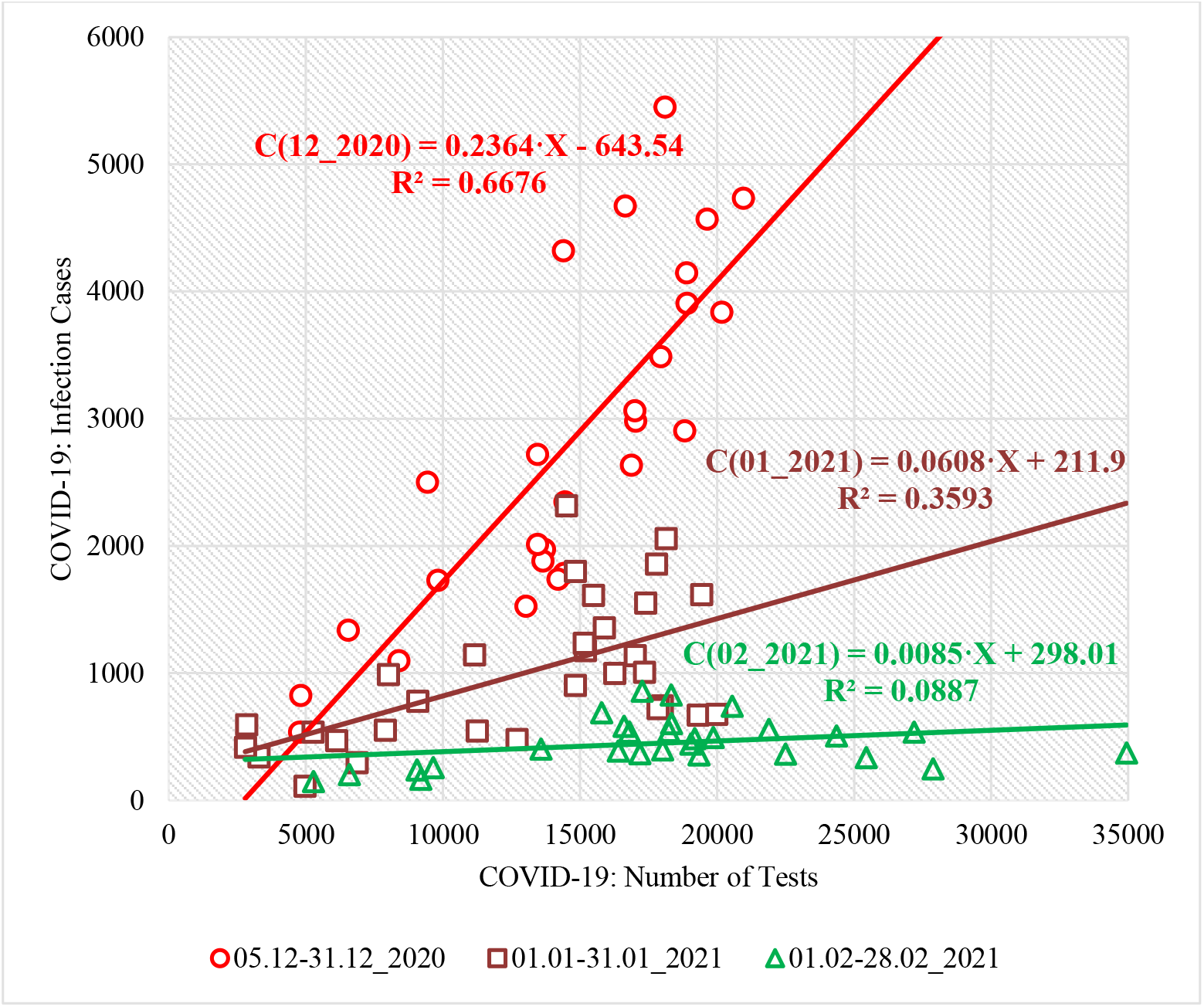
Linear correlation and regression between the confirmed cases of COVID-19 and number of tests on them performed in Georgia. C (12_2020) - 2020, December (05-31); C (01_2021) - 2021, January (01-31); C (02_2021) - 2021, February (01-28); X - Number of Tests

As follows from Fig. 15, from December 2020 to February 2021, the correlation between the confirmed cases of COVID-19 and number of tests on them significantly weakened: 2020_ December, r=0.82 (α<0.005); 2021_ January, r=0.6 (α<0.005); 2021_ February, r=0.3 (α=0.1). There are clearly positive tendencies in the spread of COVID-19.

As noted above (Fig. 11 and 12), there is some delay in the values of the time series R and D with respect to C. An estimate of the values of this delay is given below (Fig. 16).

**Fig.16.**
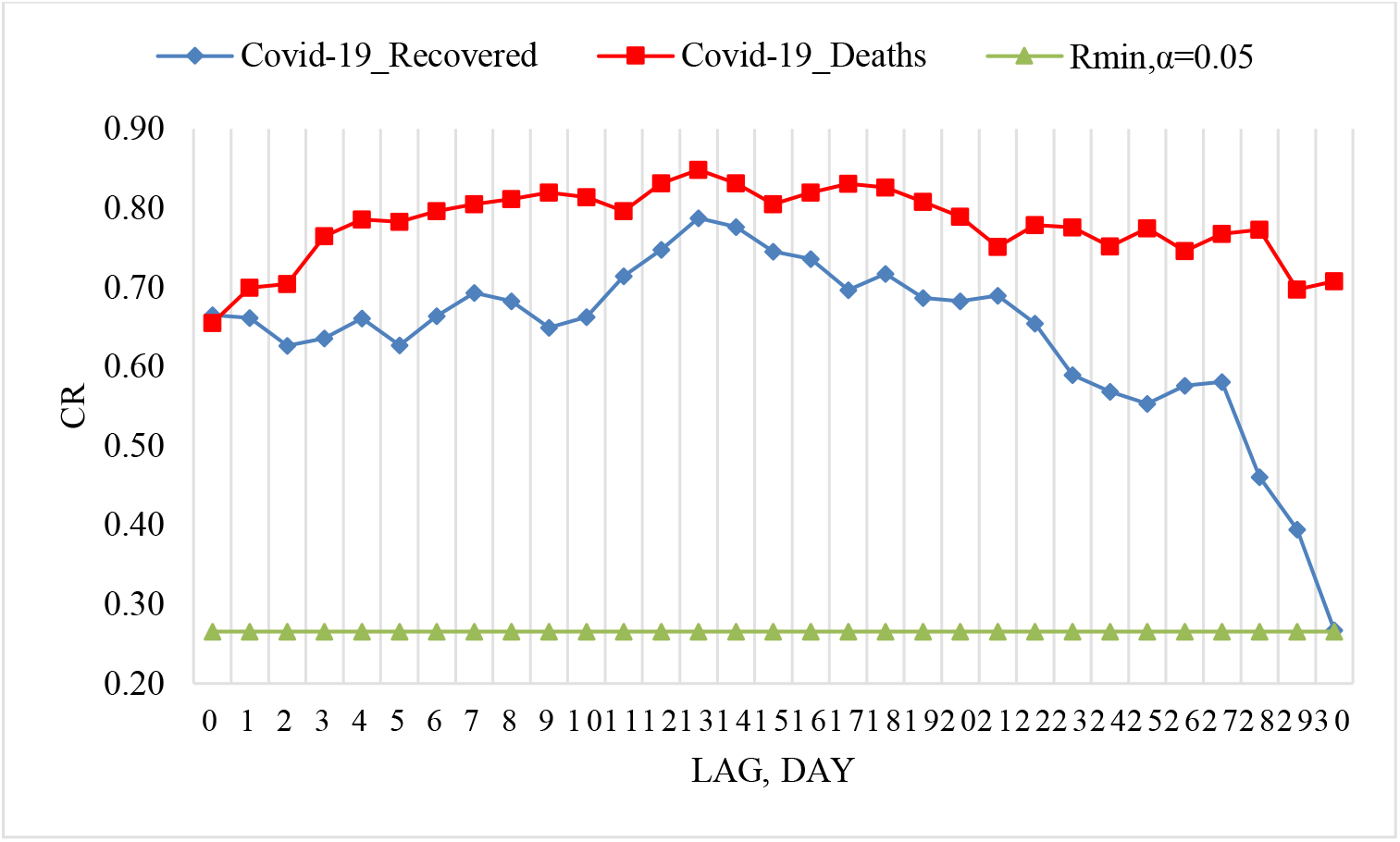
Coefficient of cross-correlations between confirmed COVID-19 cases (normed to tests number) with recovered and deaths cases in Georgia from 05.12.2020 to 28.02.2021.

Cross-correlations analysis between confirmed COVID-19 cases with recovered and deaths cases showed that the maximum effect of recovery is observed 13-14 days after infection, and deaths - after 13-14 and 17-18 days (Fig. 16). Although for fatal cases, the max of this delay are not very clearly expressed. That is, a fatal case can occur both soon enough and after a long time after infection. These effects will be refined as new data accumulates.

### 3.4 Comparison of real and calculated prognostic daily, mean decade and two weekly data related to the New Coronavirus COVID-19 pandemic

In Fig. 17-20 and Table 4 examples of comparison of real and calculated prognostic daily, mean decade and two weekly data related to the COVID-19 coronavirus pandemic are presented. Note that the results of the analysis of ten-day (decade) and two-week forecasting of the values of Č, Ď and Ĭ, information about which was regularly sent to the National Center for Disease Control & Public Health of Georgia and posted on the Facebook page https://www.facebook.com/Avtandil1948/.

**Table 4.**
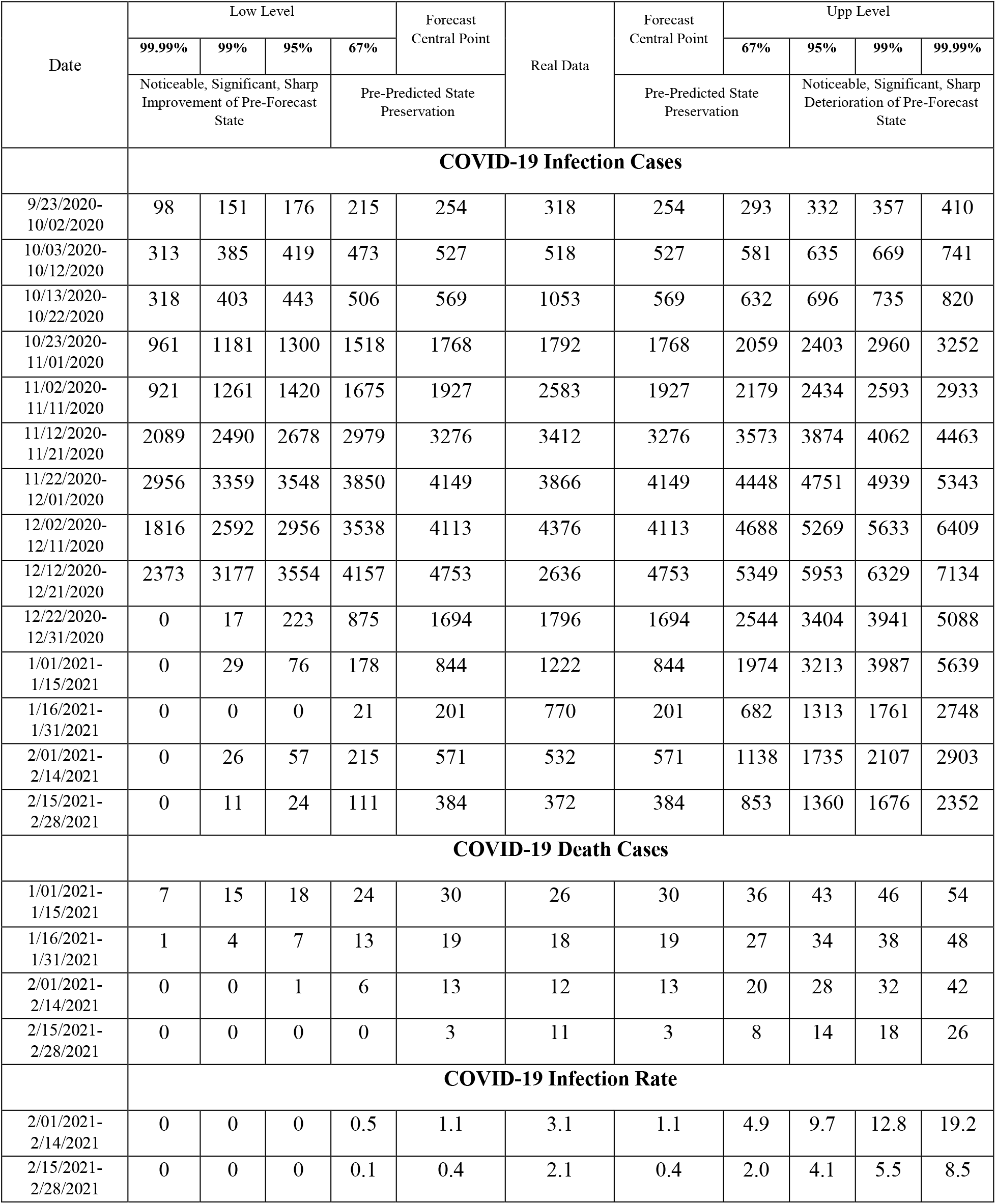
Verification of decade and two-week interval prediction of COVID-19 confirmed infection cases, deaths cases and infection rate in Georgia from 23.09.2020 to 28.02.2021.

**Fig.17.**
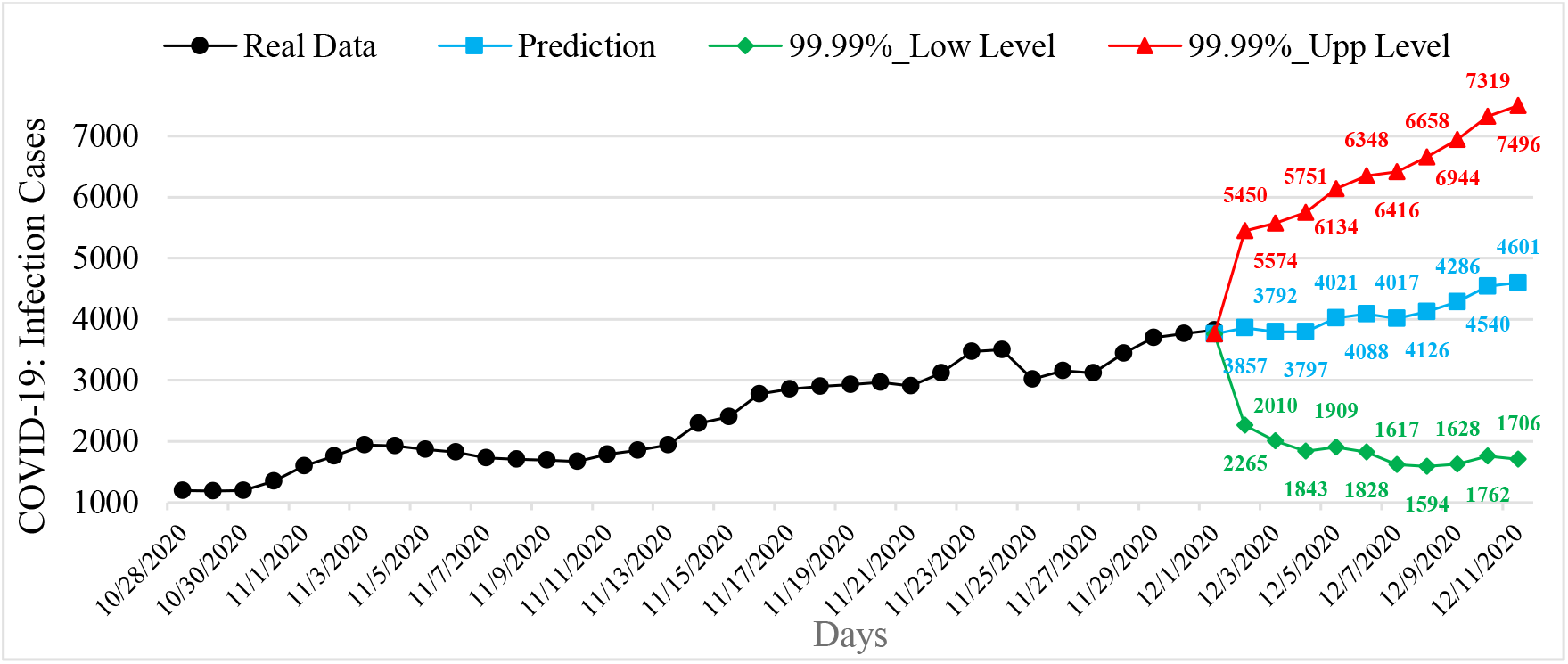
Example of interval prediction of COVID-19 infection cases in Georgia from 02.12.2020 to 11.12.2020.

**Fig.18.**
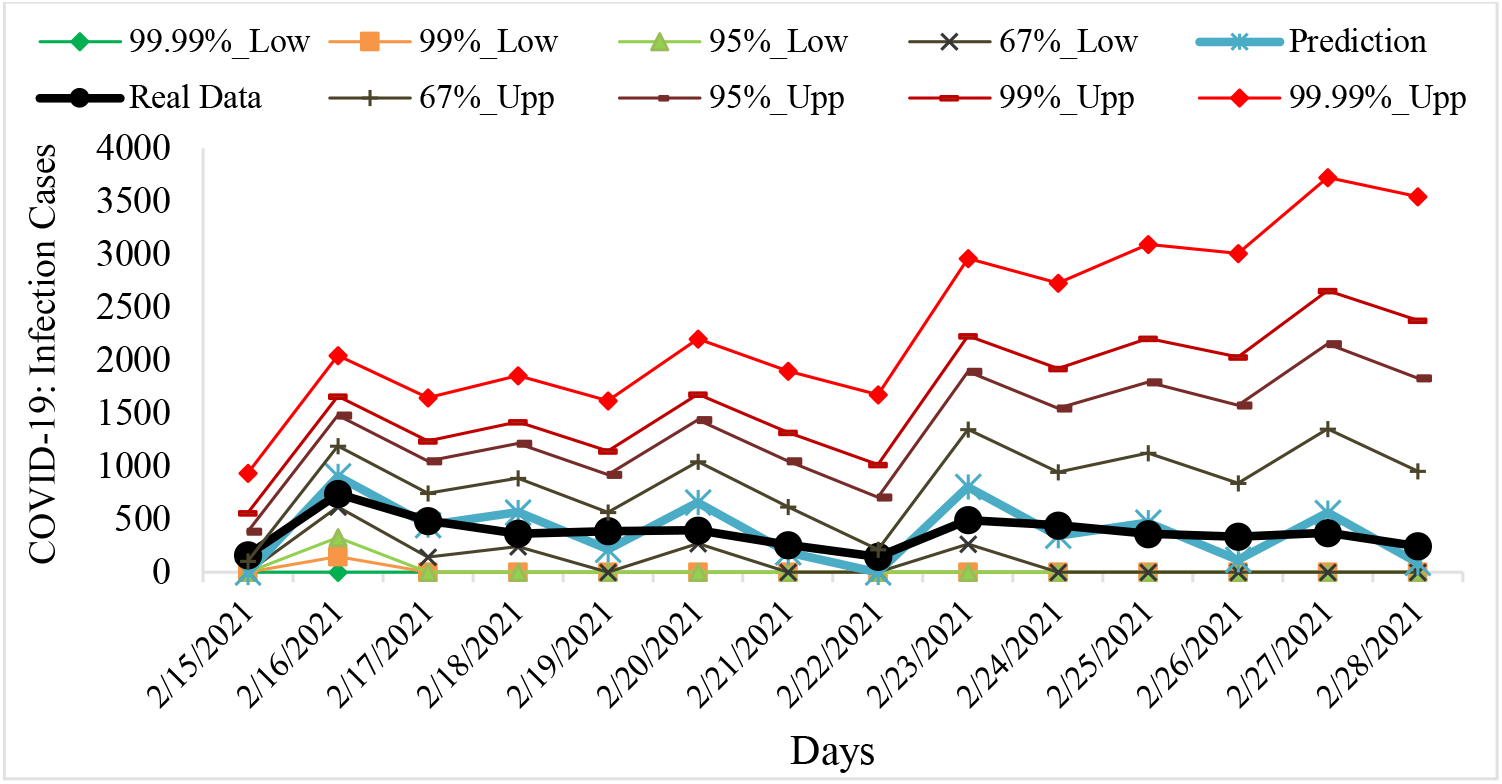
Example of verification of interval prediction of COVID-19 infection cases in Georgia from 15.02.2021 to 28.22.2021.

**Fig.19.**
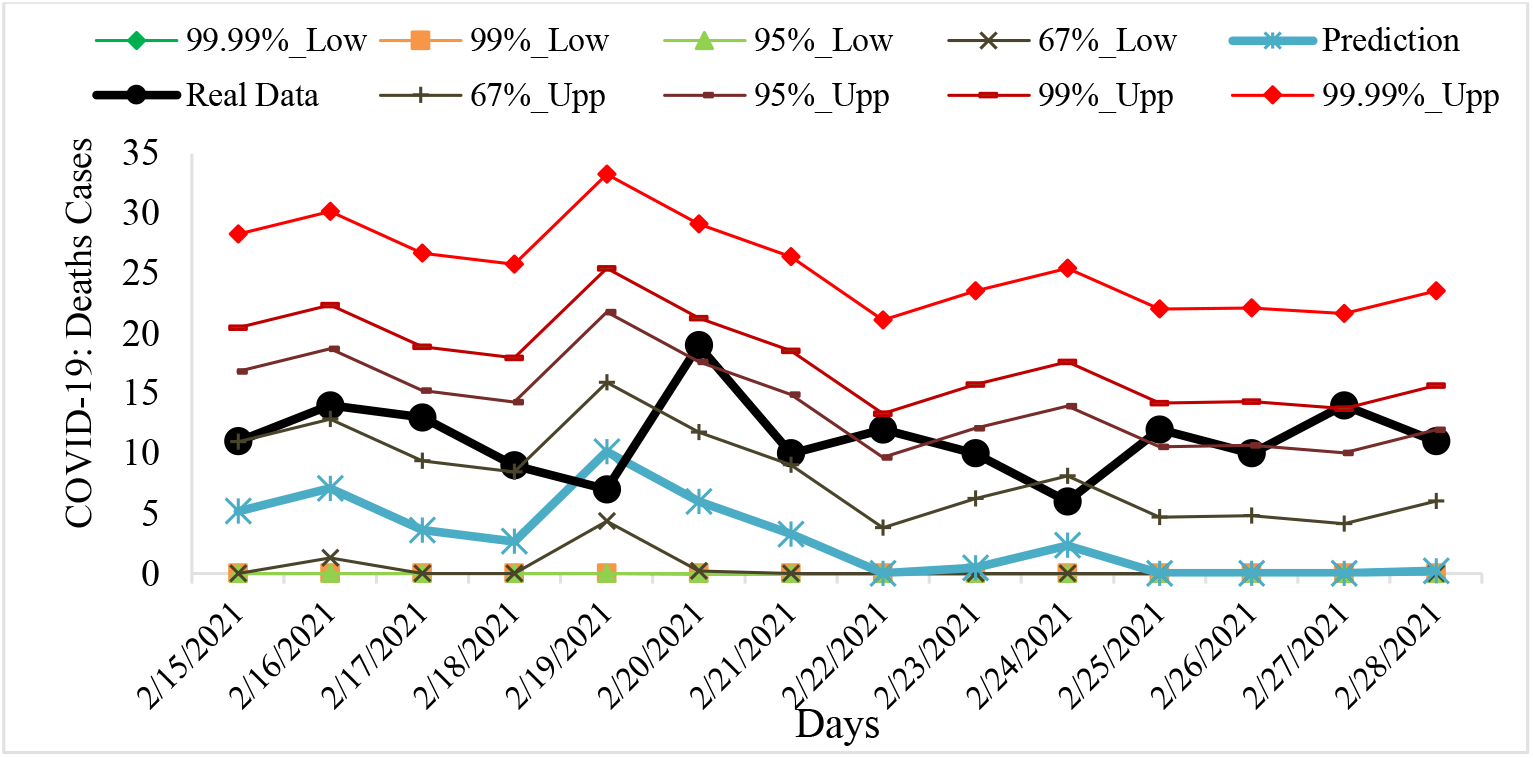
Example of verification interval prediction of COVID-19 death cases in Georgia from 15.02.2021 to 28.22.2021.

**Fig.20.**
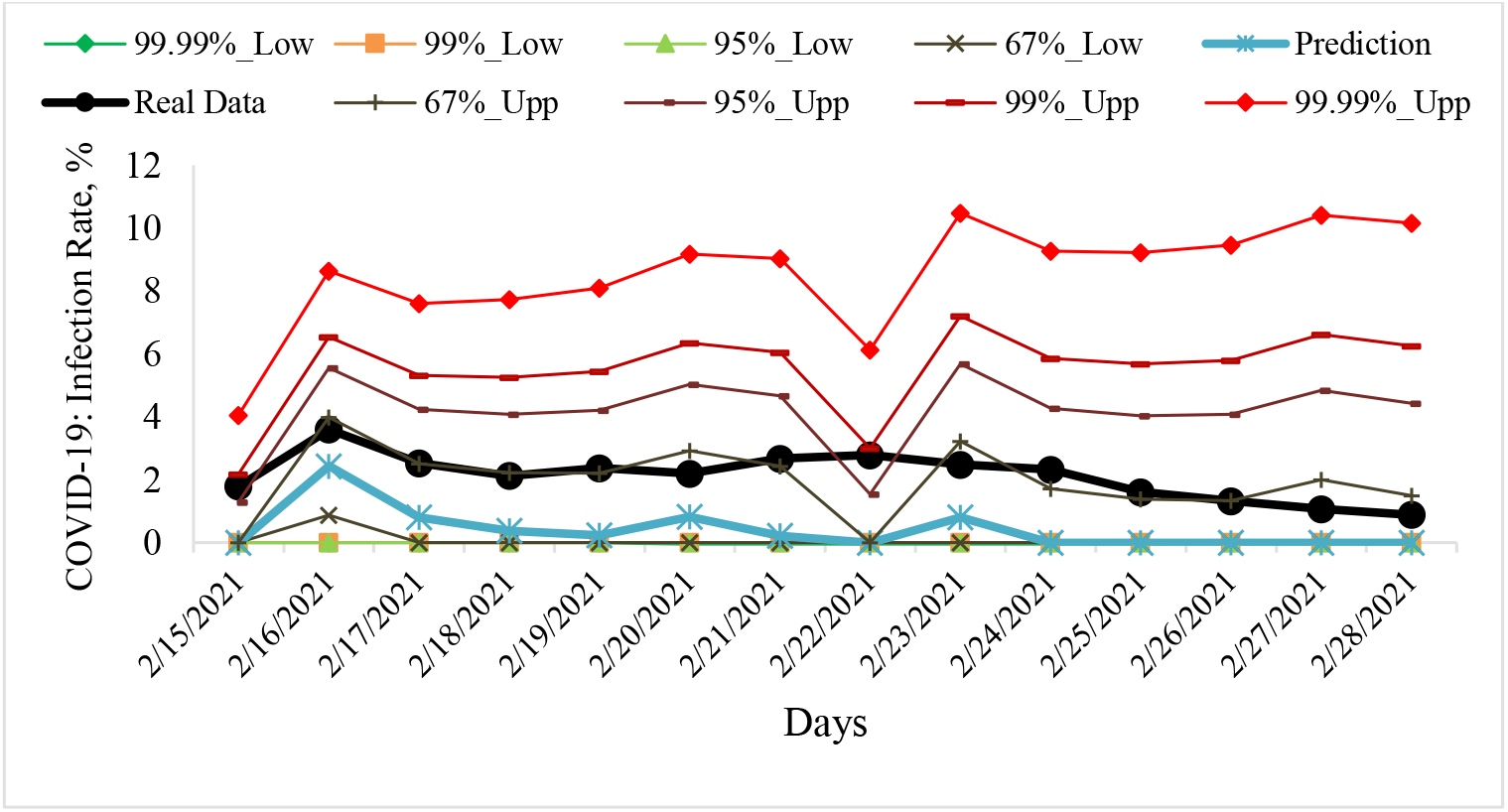
Example of verification interval prediction of COVID-19 infection rate in Georgia from 15.02.2021 to 28.22.2021.

Comparison of real and calculated predictions data of C (23.09.2020-28.02.2021), C (01.01.2021-28.02.2021) and I (01.02.2021-28.02.2021) shown that that daily, mean decade and two-week real values of C, D and I practically falls into the 67% - 99.99% confidence interval of these predicted values for the specified time periods (Fig. 17-20, data from https://www.facebook.com/Avtandil1948/, Table 4). Only the forecast of C for 13.10.2020-22.10.2020 when a nonlinear process of growth of C values was observed and its real values have exceeded 99.99% of the upper level of the confidence interval of forecast (Table 4). In this case alarming deterioration and violation of the stability of a time-series of observations was noted (according to Table 1).

For all other forecast periods (13 forecasting cases of C, 4 forecasting cases of D and 2 forecasting cases of I), the stability of real time series of observations of these parameters (period for calculating the forecast + forecast period) remained.

Noticeable deterioration of the forecast state in relation to the pre-predicted one, were observed: for C - 9/23/2020-10/02/2020 and 1/16/2021-1/31/2021; for D - 2/15/2021-2/28/2021; for I - 2/15/2021-2/28/2021. For C 11/02/2020-11/11/2020 significant deterioration of the forecast state in relation to the pre-predicted one was observed. Sharp improvement of the forecast state in relation to the pre-predicted one for C - 12/12/2020-12/21/2020 was observed. For all other cases preservation of the forecast state in relation to the pre-predicted one, were observed (Table 4).

Thus, the daily, mean decade and two-week forecasted values of C, D and I quite adequately describe the temporal changes in their real values. This was also confirmed by last verification of daily and two-week interval prediction of COVID-19 confirmed infection cases, deaths cases and infection rate in Georgia from 01.03.2021 to 26.03.2021 [https://www.facebook.com/Avtandil1948/].

## Conclusion

In the autumn - winter period of 2020, very difficult situation arose in Georgia with the course of the pandemic of the New Coronavirus COVID-19. In particular, in November-December period of 2020, Georgia eight days was rank a first in the world in terms of COVID-19 infection rate per 1 million populations. The lockdown introduced in Georgia on November 28, 2020 brought positive results. There are clearly positive tendencies in the spread of New Coronavirus COVID-19 to February 2021. The main thing is to keep them.

It is planned to continue regular similar studies for Georgia in comparison with neighboring and other countries in the future, in particular, checking the temporal representativeness of short-term interval statistical forecasting of the New Coronavirus infection COVID-19, taking into account the data on periodicity, etc.

## Data Availability

COVID-19 SARS-CoV-2 preprints from medRxiv and bioRxiv

https://stopcov.ge

